# Establishing a marine monitoring programme to assess antibiotic resistance: a case study from the Gulf Cooperation Council (GCC) region

**DOI:** 10.1101/2022.02.04.22270466

**Authors:** Edel Light, Craig Baker-Austin, Roderick M. Card, David Ryder, Mickael Teixeira Alves, Hanan A. Al-Sarawi, Khalil Hasan Abdulla, Henrik Stahl, Aliya AL-Ghabshi, Majed Al Ghoribi, Hanan H. Balkhy, Andrew Joseph, Alexandra Hughes, David W. Verner-Jeffreys, Brett P. Lyons, Will J.F. Le Quesne

## Abstract

The World Health Organization considers antimicrobial resistance as one of the most pressing global issues which poses a fundamental threat to human health, development, and security. Due to demographic and environmental factors, the marine environment of the Gulf Cooperation Council (GCC) region may be particularly susceptible to the threat of antimicrobial resistance. However, there is currently little information on the presence of AMR in the GCC marine environment to inform the design of appropriate targeted surveillance activities. The objective of this study was to develop, implement and conduct a rapid regional baseline monitoring survey of the presence of AMR in the GCC marine environment, through the analysis of seawater collected from high-risk areas across four GCC states: (Bahrain, Oman, Kuwait, and the United Arab Emirates). 560 *Escherichia coli* strains were analysed as part of this monitoring programme between December 2018 and May 2019. Multi-drug resistance (resistance to three or more structural classes of antimicrobials) was observed in 32.5% of tested isolates. High levels of reduced susceptibility to ampicillin (29.6%), nalidixic acid (27.9%), tetracycline (27.5%), sulfamethoxazole (22.5%) and trimethoprim (22.5%) were observed. Reduced susceptibility to the high priority critically important antimicrobials: azithromycin (9.3%), ceftazidime (12.7%), cefotaxime (12.7%), ciprofloxacin (44.6%), gentamicin (2.7%) and tigecycline (0.5%), was also noted. A subset of 173 isolates was whole genome sequenced, and high carriage rates of *qnrS1* (60/173) and *bla*_CTX-M-15_ (45/173) were observed, correlating with reduced susceptibility to the fluoroquinolones and third generation cephalosporins, respectively. This study is important because of the resistance patterns observed, the demonstrated utility in applying genomic-based approaches to routine microbiological monitoring, and the overall establishment of a transnational AMR surveillance framework focussed on coastal and marine environments.

## Introduction

Globally, antimicrobial resistance (AMR) has emerged as a critically important threat to both animal and human health. The reduction in effectiveness of antimicrobials used to treat pathogenic bacterial infections represents a significant global healthcare problem. Drug resistant infections are rising with estimates suggesting up to 10 million fatalities each year by 2050 due to antibiotic-resistant infections in Europe and the US alone (O’Neill, 2016), with an annual global shortfall in GDP predicted of between $1 trillion and $3.4 trillion by 2030 (World Bank, 2017). While many studies have addressed the role of AMR in clinical settings, there is growing interest in the role of the natural environment in modulating risks associated with AMR. Indeed, the growing concern over the clinical threat posed by antibiotic resistant bacteria to human health has finally turned attention to the environmental dimensions of the problem (Karkman *et al*., 2019, Wellington *et al*., 2013). Aquatic and marine environments, which receive significant inputs of pharmaceuticals, sewage waste, AMR bacteria, their associated resistance genes as well as mobile genetic elements harbouring AMR have become the focus of scientific interest (Taylor *et al*., 2011). Discharge of faecal sources into the aquatic environment, via wastewater treatment works, illegal discharges, or agricultural run-off, are key pathways for the input of antimicrobial products, resistance genes and bacteria into marine and aquatic environments (Baquero *et al*., 2008). In addition, treated or untreated sewage discharges are known to contain a diverse array of anthropogenic pollutants that have been shown to co-select for AMR (Amos *et al*., 2014, Baker-Austin *et al*., 2006). The analysis of environmental waters is being adopted as an effective method to monitor the dynamics of antibiotic-resistant pathogens (Amos *et al*., 2014), particularly given the clear route back into human populations that this environmental niche represents. It has become increasingly recognized that the ability to scrutinize the prevalence, as well as the characteristics, of resistance in environmental bacteria is critical, as part of a One Health approach, to be able to fully understand the AMR situation in clinical and community settings (Walsh, 2018).

To date, much of the data underpinning AMR risk from environmental studies has been obtained in the USA and Europe, with few published studies regarding health risks gathered elsewhere. Due to specific demographic and environmental factors the Gulf Cooperation Council (GCC) region may be particularly susceptible to the threat of AMR, with the marine and aquatic environment potentially playing a specific role in its development and propagation (Le Quesne *et al*., 2018). The Gulf is a shallow sea naturally exposed to extreme conditions of temperature and salinity due to its location, semi-enclosed nature, bathymetry and restricted circulation (Sheppard *et al*., 2010, Khatir *et al*., 2020) (Figure 1). As such, pollutants such as sewage contamination, pharmaceuticals and heavy metals entering this system may exert additional selection pressures. The region has recently undergone rapid change; the population of the GCC has increased in size by 17 million over the last 2 decades, largely attributed to an influx of foreign labor, which in many Gulf states out number nationals (Abdul Salam *et al*., 2015). This rapidly increasing and geographically diverse mixing of the GCC population, provides the potential for AMR pressures to have intensified. Furthermore, studies investigating the drivers for AMR development in the GCC suggest that there is a high rate of inappropriate antibiotic use (Al-Yamani *et al*., 2016; Balkhy *et al*., 2016; Butt *et al*., 2017) leading to recommendations calling for the implementation of regional guidelines for the management of common bacterial infections (Balkhair *et al*., 2014; Al-Yamani *et al*., 2016). Because the Gulf is also of societal and economic importance to countries bordering its waters, providing food, and drinking water (via desalination) along with supporting tourism and recreational activities, there is a clear need for a greater understanding of AMR risk in the region, particularly from an environmental context.

**Figure 1.**
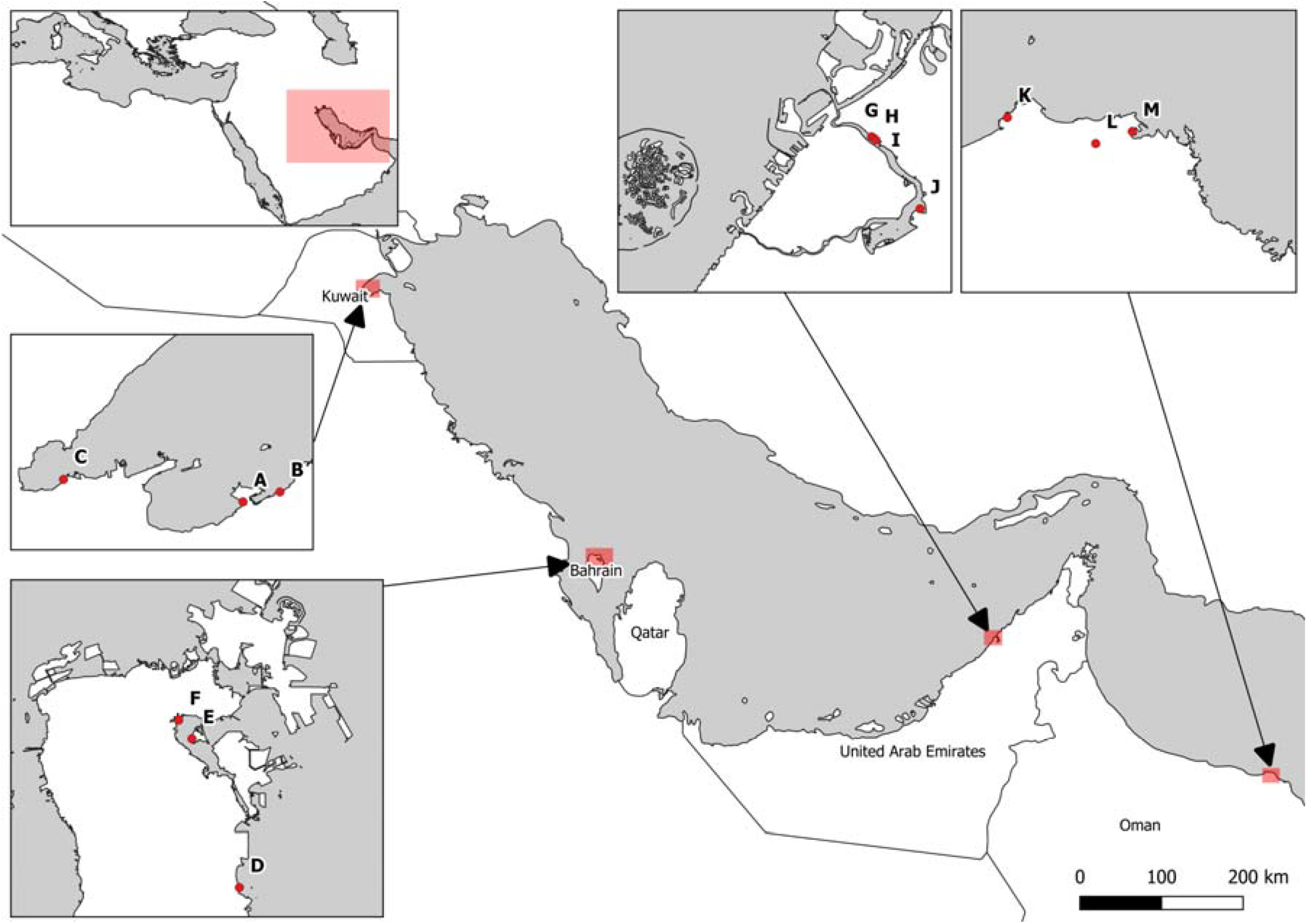
Map showing sampling locations from the different Gulf States involved in the study.

In 2018 we published a discussion article outlining a conceptual framework to more fully understand the risks associated with AMR in the marine and aquatic environment in the GCC region (Le Quesne *et al*., 2018). As part of this discussion paper, we proposed a potential methodological approach to more fully understand how such environments interact with the wider development and propagation of resistance, including a collaborative multi-national study plan for the region. We focused our investigations on *Escherichia coli*. This species is regularly used as a marker of faecal contamination from both humans and animals and has been recommended for use as an indicator for the surveillance of antimicrobial resistance (AMR) in the environment (Anjum *et al*., 2021). It is already part of established human and livestock surveillance programmes, with verified methods for its isolation and characterisation. Reference laboratories can easily implement *E. coli* in AMR environmental monitoring programmes (Anjum *et al*., 2021). As regards marine monitoring, Alves *et al*., 2014, have shown that *E. coli* can survive in coastal waters. It is relatively robust and easy to grow and there are existing antimicrobial sensitivity test (AST) protocols along with interpretive criteria for many antibiotics, including high priority critically important antimicrobials (HP-CIA, WHO 2019). Here we present the findings of a coordinated multinational monitoring program to assess the prevalence of AMR across the marine waters of the Gulf. Results from this regional baseline survey of the presence of AMR in the GCC marine environment, through the analysis of *E. coli* isolated from seawater collected from high-risk areas across four GCC states, are presented along with potential next steps for long-term implementation.

## Materials and methods

### Sample site choice and sampling plan

Representatives from the six GCC states of Bahrain, Oman, Kuwait, United Arab Emirates, Saudi Arabia, and Qatar were contacted in the Autumn of 2018 for inclusion in the study. It was agreed from the outset that National partners would collect water samples from at least three separate coastal locations, two highly impacted locations near to (any) known sewage inputs, and one sampling location in an area used for recreational activity, but close to potential sewage inputs (Figure 1). To assist in regional coordination and consistency between sampling campaigns a study plan was circulated to all partners detailing sampling methodology and initial sample processing (Supplementary File 1). Water filtration and bacterial isolation took place at laboratories in each GCC state, and isolates returned to Cefas for downstream analysis. Where project partners did not have the capacity to process samples beyond filtration, the original culture media plates were sent to Cefas laboratories for processing and analysis.

### Water sampling

At least 200 mL of water was collected from three sites in Bahrain and Kuwait and four sites in Oman and the United Arab Emirates (UAE) between December 2018 and May 2019. Two samples were taken from Al Qurum, Oman. Four samples were taken from Dubai Creek, UAE. Due to small sample sizes the data from these sites was combined giving a total of three sample sites from Oman and one from UAE. It was not possible to include samples from either Saudi Arabia or Qatar due to time constraints on the study. Water samples were homogenized and kept at < 10 °C between collection and processing. Data regarding water temperature, salinity and other pertinent observations were collected during sampling, (Supplementary File 2).

### Microbiological analysis

For the detection and isolation of *E. coli* through membrane filtration, the following procedure was adopted: 10 mL of sample was serially diluted in 1/4 strength Ringers Solution (10^0^, 10^−1^, 10^−2^, 10^−3^, 10^−4^, and 10^−5^) and filtered through 47 mm 0.45 μm nitrocellulose membrane; the membrane was placed on a plate of Tryptone Bile X-Glucuronide Medium, TBX agar (Oxoid), and incubated at 30 °C for 4 hours followed by 44 °C for between 17 and 21.5 hours, one laboratory omitted the 30 °C incubation step; finally, the number of positive (blue-green) colonies on each plate was counted (Vergine *et al*., 2017) and total *E. coli* enumerated. Blue-green colonies were picked off the plates using sterile loops, inoculated onto nutrient agar slants (Oxoid), and incubated overnight at 44 °C. The slants were stored at 2 - 8 °C before shipping to Cefas (Weymouth, UK) for further microbiological and AMR analyses. The combination of culture medium and incubation temperature was deliberately chosen, for health and safety reasons, to exclude the isolation of toxigenic *E. coli*. The study plan recommended 50 *E. coli* isolates from each location. However, that was not always possible. In locations with a low *E. coli* count all blue-green colonies were selected. In areas with a high *E. coli* count discreet colonies were picked *ad hoc* from the plates. Total numbers were made up by utilising more colonies from areas with a high *E. coli* count. Differences in number between locations were accounted for during statistical analysis.

### Antimicrobial susceptibility testing

Upon receipt at Cefas, isolates were removed from slopes onto TBX plates and incubated overnight at 44 °C. Cultures were purified using further subculture steps, before storage on Protect™ beads at −80 °C. Some cultures could not be revived. Numbers of cultures refer to those which were viable and not total isolates received. The control strain *E. coli* ATCC 25922 was used throughout. All *E. coli* isolates, were tested for phenotypic resistance by broth microdilution using the Trek Sensititre system (Thermofisher), following manufacturer’s instructions. All isolates were tested on the EU surveillance EUVSEC plate for susceptibility towards 14 antibiotics: Ampicillin, Azithromycin, Cefotaxime, Ceftazidime, Chloramphenicol, Ciprofloxacin, Colistin, Gentamicin, Meropenem, Nalidixic acid, Sulfamethoxazole, Tetracycline, Tigecycline and Trimethoprim. The European Committee on Antimicrobial Susceptibility Testing (EUCAST) epidemiological cut-off values (ECOFF) were used for interpretation of wild type vs. non-wild type (WT/NWT) where wild type bacteria possess no acquired resistance mechanisms, and non-wild type bacteria have an acquired resistance mechanism (Schwarz *et al*., 2010). All isolates showing reduced susceptibility to Cefotaxime and/or Ceftazidime (3^rd^ generation cephalosporins) were tested on the EU surveillance extended spectrum beta lactamase (ESBL) EUVSEC2 plate to determine susceptibility to: Cefoxitin, Ertapenem, Imipenem, Meropenem, Cefotaxime, Ceftazidime, Cefepime, Cefotaxime/Clavulanic acid, Ceftazidime/Clavulanic acid, and Temocillin. This plate is used to elicit antibiotic resistance phenotypes such as: Extended spectrum beta lactamase ESBL, ampicillinase C AmpC, ESBL+AmpC and putative carbapenamase producers, using the criteria published in the technical specifications on harmonised monitoring of antimicrobial resistance in zoonotic and indicator bacteria from food-producing animals and food (EFSA, 2019). Multidrug resistance was defined as any isolate showing the NWT phenotype to three or more classes of antibiotic, EFSA 2020. Throughout the paper any mention of ‘resistance’ as in multidrug resistance MDR, phenotypic resistance etc. refers to the non-wild type phenotype, which is not necessarily synonymous with clinical resistance.

### Statistics

Pearson’s correlation coefficients were calculated for identifying associations between countries and locations. To assess differences between locations within countries a linear mixed model was conducted considering paired measurements between individual antibiotics, MDR, ESBL and AmpC. Normality of the residuals was verified. A pairwise posthoc test was conducted for estimating locations that differed from each other.

Cohen’s Kappa test was used to compare MIC with the presence of resistance genes for 8 of the antimicrobials. We used the interpretation as described by Stubberfield *et al*., 2019 where a k value of 0 indicates a test that agrees as well as could be expected, compared to a k of 1 which indicates complete agreement. A result of k > 0.900 was interpreted as almost perfect agreement, k 0.800 – 0.899 as strong agreement, k 0.600 – 0.799 as moderate agreement, k 0.400 – 0.599 as weak agreement and k 0.200 – 0.399 as minimal agreement.

### Whole genome sequencing

A subset of 173 isolates was chosen for whole genome sequencing, to provide insight into the genetic diversity of the isolates and the AMR resistance determinants they harboured. The selection was made to encompass isolates with resistance to HP CIAs and/or having an MDR phenotype. Isolates from all four countries were included in the selection. Briefly, genomic DNA was extracted from 173 strains following the QIAamp DNA Mini kit protocol. Libraries were prepared with the Nextera XT DNA sample preparation kit from Illumina and sequenced using two lanes on a NovaSeq 6000, with 150 cycles of paired-end sequencing.

### Quality Control

FastQC version 0.11.8 and MultiQC version 1.7 were used for assessing the quality of raw sequencing data (Andrews *et al*., 2016). KAT version 2.4.1 was used to compare 27bp kmers from sequences which were assembled, as described below, and raw sequencing data (Mapleson *et al*., 2017). Any samples with two or more coverage peaks or which had a large number of distinct, high coverage kmers which aligned against multiple, different parts of the assembly were identified as having a large degree of heterogeneity, presumed due to inclusion of multiple bacterial isolates during library preparation of a single sample, and were therefore excluded. Isolates were also excluded due to poor coverage or assembly of sequencing data, as assessed using the Nullarbor pipeline.

### Analysis of Bacterial Isolates using Nullarbor

Version 2 of the Nullarbor pipeline was used to generate a report on each bacterial isolate (Seemann *et al*., 2020), which includes steps for cleaning reads, species identification, *de novo* assembly, multi-locus sequence typing and calling Single Nucleotide Polymorphisms (SNPs). Removal of adaptors and low-quality bases or reads was carried out using version 0.39 of Trimmomatic (Bolger *et al*., 2014). Each bacterial isolate was assigned to the closest matching species using version 2.0.8 of the kraken2 software (Wood *et al*., 2019), which used version 1 of the minikraken2 database, as released in April 2019, as a reference. Reads were assembled into a draft genome using SKESA version 2.3.0 (Souvorov *et al*., 2018). The sequence type of each bacterial isolate was identified using version 2.16.1 of the MLST software (Seemann, 2020a), parameterised to search against the *E. coli* sequence typing scheme. The MLST software in turn makes use of the PubMLST website (https://pubmlst.org/) developed for microbial MLST designations (Jolley and Maiden, 2010). Snippy version 4.3.6 was used to call SNPs (Seemann, 2020b), with the published genome of *E. coli* strain K-12 sub-strain MG1655 being used as a reference (accession number GCA_000005845.2). The core SNPs, as identified by snippy, were used to infer phylogeny, using version 1.6.10 of IQ-tree (Nguyen *et al*., 2015), which was parameterised to use the maximum-likelihood general time reversible model with unequal rates and base frequencies as well as four rate categories and the fast tree search mode.

### Identification of resistance genes using ResFinder

Version 4.0.0 of the ResFinder software was downloaded and installed on the 11^th^ of November 2020, along with the latest version of the resfinder and pointfinder database (Bortolaia *et al*., 2020; Zankari *et al*., 2013). Results where coverage and identity scores were equal to one were reported as normal. Genes identified in isolates with a more fragmented genome assembly, where there was incomplete coverage of the gene coding region, were only reported as belonging to a specific gene family, rather than being reported as a specific allele, and were described as *‘partial sequences’*. Complete gene sequences, which aligned against a reference, but with a lower identity score of 0.95 and 0.99, were described as *‘novel variants’*. In either case, sequences were aligned against reference sequences from the ResFinder database to confirm the result, using MAFFT version 7.471 and UGENE v37.0 (Katoh *et al*., 2013; Okonechnikov *et al*., 2012).

### Visualisation of Results

The R statistical programming language (v3.5.1) was used for processing results from antibiotic testing (R Core Team, 2018), along with the readxl (v1.3.0) for importing Excel spreadsheets (Wickham *et al*., 2019), ggplot (v3.1.0) for plotting data (Wickham, 2009), dplyr (v0.8.0.1) for creating summary statistics (Wickham *et al*., 2019), stringr (v1.3.1) for running various regular expressions (Wickham, 2018), tidyr (v0.8.3) for reshaping tables from the wide to long convention (Wickham and Lionel, 2019), nlme for fitting the linear mixed model to the data (Pinheiro *et al*., 2020) and multcomp for conducting pairwise comparisons (Hothorn *et al*., 2008). Phylogenetic inferences, along with phylogenetic data and information on presence and absence of specific resistance genes was then plotted using version 5.5 of the ITOL webservice (Letunic and Bork, 2019).

## Results

### Microbiological

A total of 560 *E. coli* isolates was analysed for this monitoring programme, sampled between December 2018 and May 2019, and obtained from Kuwait (n=223), Bahrain (n=156), Oman (n=152) and UAE (n=29) (Supplementary File 2). Isolates fully susceptible to all 14 antimicrobials tested comprised 47.5% (n=266), whereas 30.5% (n=171) of isolates were multidrug resistant. The most commonly observed resistance phenotypes were: ciprofloxacin (44.6%), ampicillin (29.6%), nalidixic acid (27.9%), tetracycline (27.5%), trimethoprim (25.5%) and sulfamethoxazole (22.5%). Resistance to chloramphenicol was 6.6%. Resistance to the third generation cephalosporins cefotaxime and ceftazidime was at 12.7%; 8.8% of which were classed as having the ESBL phenotype, 2.0% with having the AmpC phenotype and 1.4% expressing both ESBL and AmpC phenotypes. The proportion of isolates showing resistance to other HP-CIAs was as follows: azithromycin (9.3%), gentamicin (2.7%), colistin (0.5%) and tigecycline (0.5%). No isolates showed resistance to meropenem. The overall phenotypic results, and those for the individual sampling sites, are indicated in Table 1.

**Table 1.**
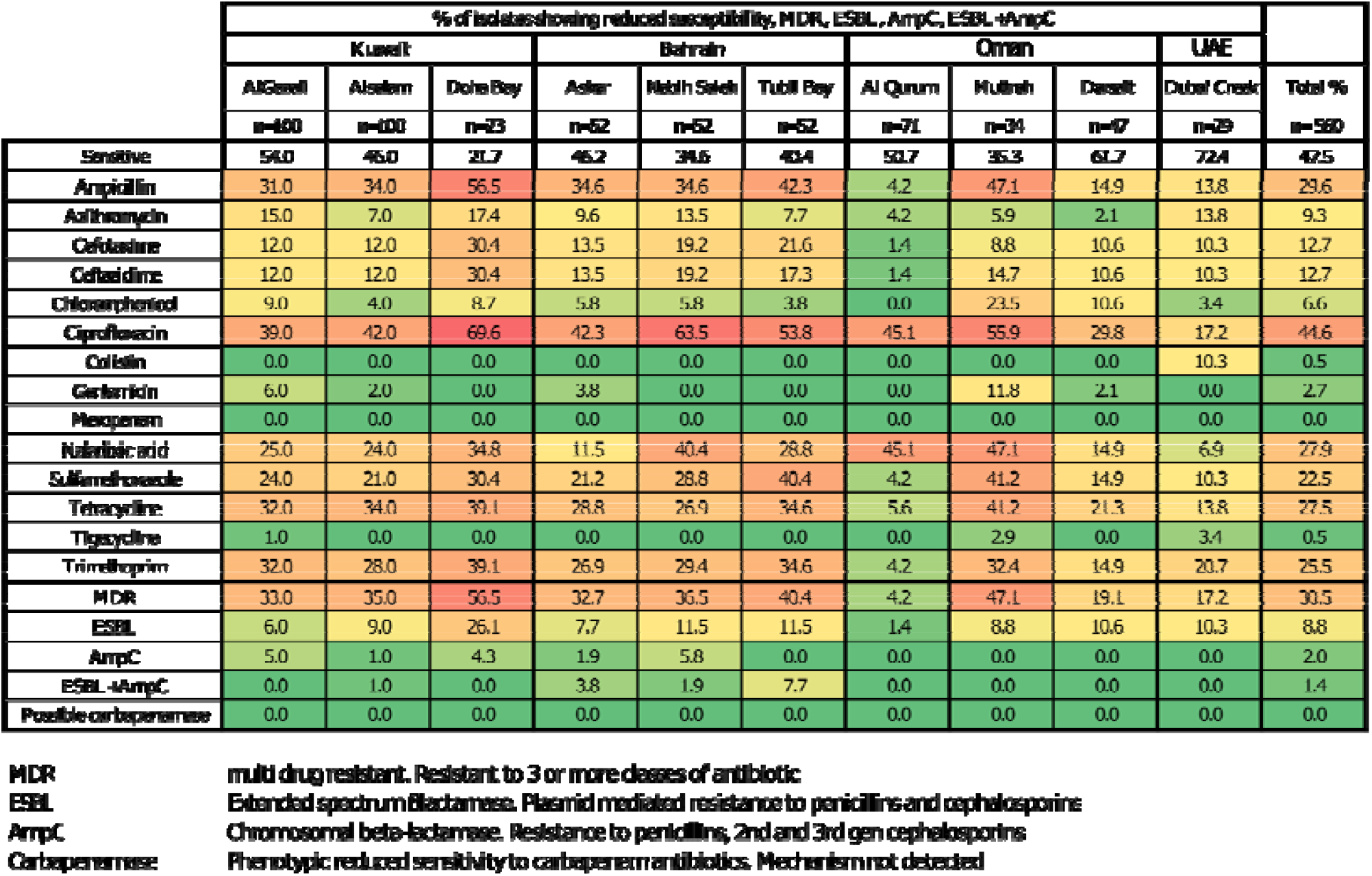
Heatmap depicting percentage resistance, MDR, and ESBL, AmpC phenotypes of analysed *E. coli* isolates. Raw MIC data can be seen in Supplementary file 3.

The statistical analysis looked for significant differences at the 5% level between levels of phenotypic resistance to antibiotics, ESBL and AmpC phenotpyes, and MDR. These were compared firstly between countries, and secondly between the different locations within each country. When looking at resistance there was a strong positive association between Kuwait, Bahrain and Muttrah in Oman (r>0.85). No statistical difference in levels of MDR, ESBL or AmpC was found between countries. Within Kuwait, Doha Bay had significantly higher levels of resistance to antibiotics than either AlGazali (p=0.0002) or AlSalam (p=1.10^−4^). Doha Bay also had significantly higher numbers of ESBL producers than AlGazali (p=0.007). In Bahrain both Nabih Saleh and Tubli Bay had significantly higher levels of resistance than Askar (p=0.014 and p=0.044; respectively). In Oman, Muttrah had significantly higher levels of resistance than either AlQurum (p=1.10^−4^) or Darsait (p=1.10^−4^). All three locations, Muttrah, Darsait and AlQurum were significantly different from each other as regards numbers of multidrug resistant isolates with Muttrah having the highest level followed by Darsait. There were very low numbers of isolates returned from UAE. These were sampled from 4 areas sited geographically close together. They were regarded as a single sample and as such there is no within country comparison for UAE.

### Whole Genome Sequencing

#### Genomic analyses

173 *E. coli* were sequenced. They were selected according to their phenotypic resistance profiles, with a focus on isolates with multidrug resistance and/or resistance to HP CIAs. The median depth of sequencing coverage was 306x and isolates aligned against at least eighty percent of the reference genome *Escherichia coli* K-12 (GCA_000005845.2). Individual assembly data is available in Supplementary file 4. The raw and assembled sequencing data have been submitted to NCBI and archived under BioProject accession number PRJNA691754.

#### Genotype v phenotype

Cohen’s kappa test was used to compare MIC with the presence of resistance genes for ampicillin, cefotaxime, ceftazidime, gentamicin, azithromycin, sulfamethoxazole-trimethoprim, ciprofloxacin and tetracycline, and for the ESBL and AmpC phenotypes. There was almost perfect agreement (k>0.900) between MIC and associated resistance genes for ampicillin, tetracycline, azithromycin, ciprofloxacin, and the AmpC phenotype. There was strong agreement (k 0.800 – 0.899) for gentamicin, ceftazidime, cefotaxime, and sulfamethoxazole-trimethoprim, with moderate agreement (k 0.600 – 0.799) for the ESBL phenotype. The frequency gene observations used to make these predictions are listed in Supplementary file 5 and the positive and negative predictive values and kappa values are listed in Supplementary File 6.

#### Gene prevalence

Of the 173 sequenced isolates 39.9% did not have any resistance genes detected, while 45.1% carried resistance genes to three or more classes of antibiotic. The most prevalent genes found were *sul* (47.4%), *tet* (45.1%), *qnr* (41.6%), *dfr* (38.7%), *bla*_TEM-1_ (37.0%), *bla*_CTXM_ (26.6%), and *mph* (15.6%). No resistance genes were detected for the HP-CIA antibiotics colistin, meropenem or tigecycline (Supplementary File 5).

#### Resistance to the β-lactam antibiotics

Resistance to the β-lactam antibiotics including ampicillin and the third generation cephalosporins, cefotaxime and ceftazidime, was widespread involving several different genes. The *bla*_TEM-1_ gene conferring resistance to ampicillin was present in 37.1% of the sequenced isolates. The *bla*_DHA-1_, and *bla*_CMY_ variants, consistent with the AmpC phenotype, were found at 6.4% and 1.8% respectively. There was a very high prevalence of the ESBL gene *bla*_CTXM-15_ (25.4%), and to a lesser extent the other ESBL genes *bla*_CTXM-14b_ (0.6%), *bla*_CTXM-55_ (0.6%), and *bla*_SHV-12_ (1.2%). Figures 2(a) and 2(b) show the relationship between isolate location, sequence type, phenotypic resistance to the β-lactam antibiotics and the presence of genes associated with resistance to ampicillin and the ESBL and AmpC phenotypes.

**Figures 2a and Figure 2b.**
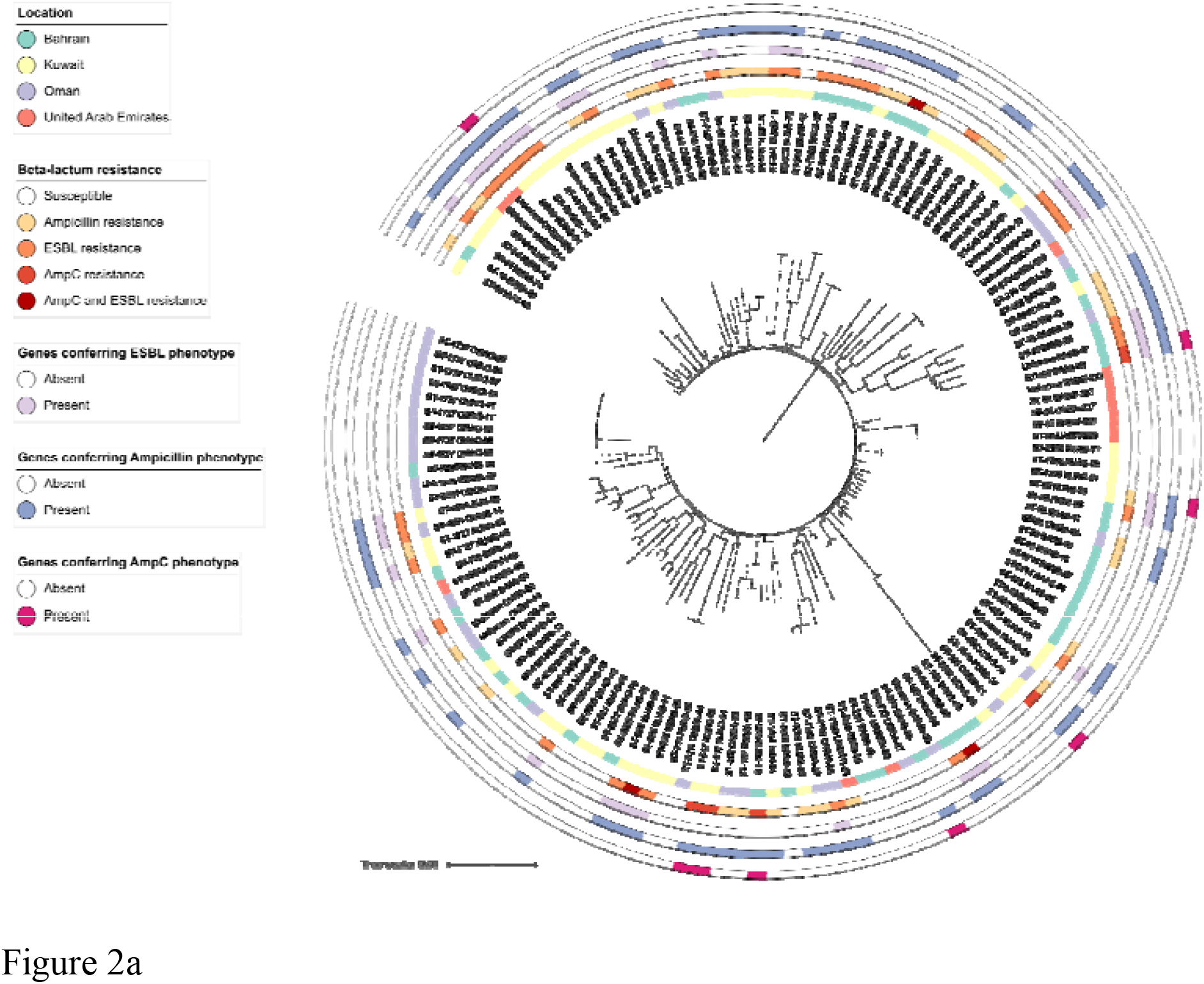
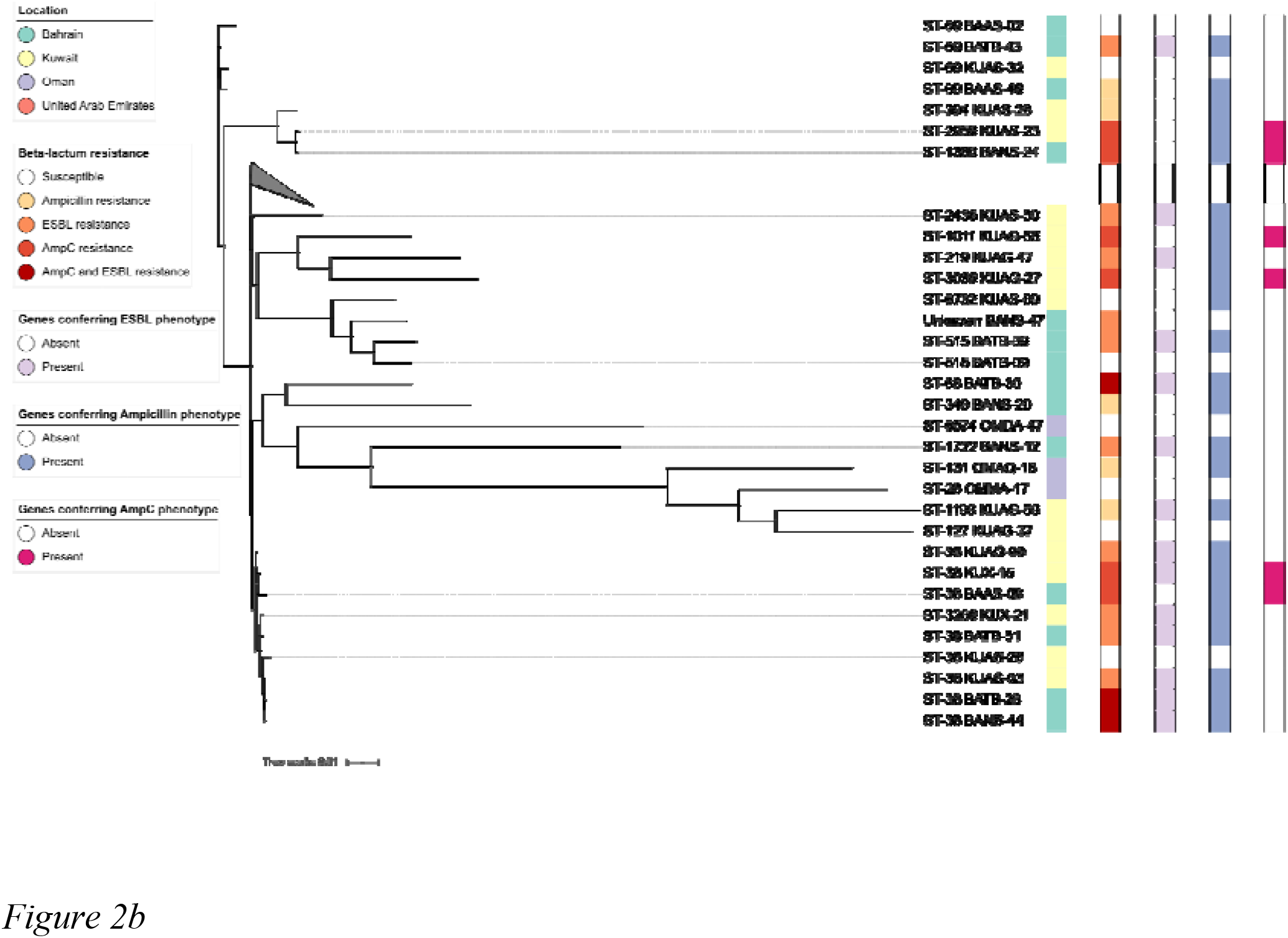
Phylogenetic trees showing isolates which are separated by an average branch distance of (a) less and (b) greater than 0.01. The beta-lactam resistance phenotype observed during phenotypic testing is included, along with any genes thought to contribute to the ESBL (*bla*_SHV-12_, *bla*_CTX-M-14b_, *bla*_CTX-M-15_, *bla*_CTX-M-55_), AmpC (*bla*_CMY-2_, *bla*_DHA-1_) and Ampicillin (*bla*_TEM-1A_, *bla*_TEM-1B_, *bla*_TEM-1C_) resistance phenotypes.

### Resistance to the quinolones

There were very high levels of resistance to the quinolones spread throughout the region. Of 246 isolates showing phenotypic resistance 103 were sequenced. There were two distinct MIC peaks for ciprofloxacin corresponding to 0.25 mg L^-1^ and >8 mg L^-1^. There were corresponding high levels of the *qnrS* genes: *qnrS*1 (56), *qnrS*13 (2), *qnrB*4 (6), *qnrS*1 with *qnrB*4 (1), *qnrB*7 (1). These tended to correlate to the lower MIC peak of 0.06<Cip≤2 mg L^-1^. Chromosomal mutations in the *gyrA, parC* and *parE* genes were noted more with the higher MIC peak 8<Cip≥32, particularly in the absence of the *qnr* genes.

### Sequence type analysis

A diverse range of sequence types was identified throughout the region. We have highlighted those associated with clinical disease, also clones within the different sequence types which were MDR and both expressed and contained genes conferring reduced susceptibility to the third generation cephalosporins. The relatedness criteria for clonality in *E. coli* was a SNP distance of ≤10 (Schürch *et al*., 2018). The most prevalent sequence types (STs) were; ST1737 (9), ST10 (8), ST38 (8) and ST155 (8). All ST1737 isolates came from the same location in Oman and had a SNP distance of 0. We believe that this was method related and the colonies stemmed from an original *E. coli* isolate. ST34, ST131, ST295, and ST746 were present in small numbers. These sequence types, including ST10, ST38 and ST155, are frequently recovered from food and human intestinal samples and have strong associations with uropathogenic and diarrhoeal disease. In particular, ST10, ST38 and ST155 isolates are found commonly in the gut of humans and food animals. They have an intrinsic ability to acquire AMR genes which may lead them to play a crucial role as a reservoir of these genes (Matamoros *et al*., 2017). Seven of the ST10 isolates were MDR and 5 carried the *bla*_CTXM-15_ gene. Seven of the ST38 isolates were MDR and 6 carried the *bla*_CTXM-15_ gene. Six of the ST155 isolates were MDR with 5 carrying either *bla*_CTXM-15_ or *bla*_CMY-2_ genes. A clone consisting of 3 isolates from Oman were MDR and carried the *bla*_CTXM-15_ gene. This clone did not have an identified ST. Fourteen clonal groups were found. Eleven involved samples taken from the same locations at the same time. The three clonal groups which contained isolates from different locations all came from Bahrain. One location was close to a wastewater outlet whilst the other was about 2km away at the other end of a bay.

## Discussion

The WHO has clearly outlined in its Global Action Plan that it is crucial that countries adopt integrated surveillance programs for monitoring antibiotic resistance. As such many countries are now implementing national surveillance programs, which cover both clinical and veterinary settings. However, few such schemes appear to exist for monitoring AMR in environmental or aquatic settings (Marano *et al*., 2020). The WHO have developed and recommended the Tricycle Programme as a mechanism to collect harmonised data on prevalence of ESBL producing *E. coli* from different sectors, including the environment (Matheu *et al*., 2017). Our study builds on this initiative to provide further phenotypic and genetic information on *E coli* recovered from marine waters in the Gulf, including whether they are ESBL producers. This data can be used to better assess transmission pathways, likely risks posed by new and emerging resistances, and better monitor the effectiveness of intervention strategies.

There are challenges associated with setting up and running transnational surveillance programs. For example, in the study presented here there were some logistical problems in isolating a representative library of *E. coli* strains from all sites and across all sampling periods. The isolates screened here probably represent different industrial, domestic, and agricultural sources encompassing diverse mammalian and avian hosts, as such it is not possible to make definitive assertions regarding the sources of observed resistances. Also, it should be noted that the study presented here is limited in terms of size and scope and represents a proof of concept regarding the application of environmental surveillance to study AMR. Irrespective, the success of such a short “snapshot” surveillance effort cannot be understated and is undoubtedly important in the global effort to tackle AMR.

The AMR surveillance programme outlined here is both important and notable for several reasons. Firstly, the initial results generated from this study show that a high proportion of the *E. coli* isolates recovered from marine waters across the region were multi-drug resistant and many showed reduced susceptibility to HP-CIAs. In this regard, the data presented here mirrors findings that have been reported from both clinical and environmental studies in the region (Al-Zarouni *et al*., 2008, Balkhy *et al*., 2016, Khan *et al*., 2020), outlining high levels of resistance in analysed strains. Of note, multi-drug resistance (resistance to 3 or more structural classes of antimicrobials) was observed in 30.5% of tested *E. coli* strains isolated in this study. High levels of reduced susceptibility to a range of structurally diverse agents, including ampicillin (29.6%), nalidixic acid (27.9%), tetracycline (27.5%), sulfamethoxazole (22.5%) and trimethoprim (25.5%) were observed (Table 1). Reduced susceptibility to the beta lactam antibiotics was widespread in the strains analysed, and across all countries. Worryingly, reduced susceptibility to HP-CIAs, including ciprofloxacin (44.6%), the third generation cephalosporins cefotaxime (12.7%) and ceftazidime (12.7%), tigecycline (0.5%) and colistin (0.5%) was also observed. Perhaps most significantly, reduced susceptibility to ciprofloxacin was detected in over 40% of tested *E. coli* isolates (Table 1). This matches previous data observed for marine isolates reported from Kuwait (Al-Sarawi *et al*., 2018). Previous meta-analysis studies have shown high level resistance to ciprofloxacin in environmental and food-derived *E. coli* strains, but at generally lower levels than observed here (14.4%, range 5.4-33.4%). However, high ciprofloxacin resistance (∼52%) has previously been described in studies of *E. coli* isolated from urine samples and poultry carcasses in Iran (Pormohammad *et al*., 2019). Identification of reduced susceptibility to colistin and tigecycline in environmental *E. coli* strains is of some concern, given that these are HP-CIA (WHO 2019). Although the levels were low (0.5%) there remains the potential for these resistances to be present at higher frequencies given the limited scope of this monitoring study. Due to the labile nature of tigecycline it is not always easy to generate an accurate MIC. All isolates with an MIC above the ECOFF were repeated. Two of these were one dilution above the ECOFF and the other two dilutions above. As tigecycline is considered a drug of last resort to treat carbapenem-resistant infections, the detection of such resistances in environmental strains is particularly noteworthy.

Secondly, as part of this surveillance programme we chose to whole genome sequence a subset of isolates (173/560 strains, ∼30% of the total library). This sequencing effort revealed an expected diversity of resistance genes as well as helped identify potentially clinically relevant sequence types (STs). Of note, high carriage rates of *qnrS1* (60/173 sequenced isolates), *bla*_CTX-M-15_ (45/173) and *bla*_DHA-1_ (11/173) were observed, correlating with reduced susceptibility to the fluoroquinolones and third generation cephalosporins, respectively. These latter genes are typically carried on plasmids and are both associated with extended spectrum cephalosporin resistance in a range of clinically important Gram-negative bacteria, including *E. coli*. Plasmid mediated resistance is particularly relevant to the persistence and spread of AMR both within and between species in the aquatic environment. Previous studies have shown that both genes are circulating in hospital and community (Peterson and Bonomo, 2005). They are also evident in pathogens in the region (Khan *et al*., 2020). Their presence at such high levels in the aquatic environment is concerning. One possible explanation could be the level of sewage pollution. Many of our samples were taken close to sewage outlets with ‘obvious pollution’ described as present at the time of sampling. Despite finding no statistical difference in the levels of resistance and MDR between countries there were some differences between locations within countries, with significantly higher levels found in busy harbours and close to sewage outlets compared to recreational beaches. Interestingly, while the rates of both ESBL and carbapenamase producing *E. coli* are increasing in clinical settings and in the community in the GCC (Zowawi *et al*., 2020) we only detected ESBL *E. coli*. We did not detect resistance to meropenem or the presence of *bla*_KPC_, *bla*_IMP_, *bla*_NDM_, *bla*_SPM_, *bla*_VIM_, or *bla*_OXA-48_. Despite seeing low level phenotypic resistance to both colistin and tigecycline we did not detect either the *mcr* or *tetX* genes associated with plasmid mediated resistance to these CIAs.

The genetic basis of resistance to front-line clinically relevant drugs is the focus of current work on several isolates, especially those demonstrating reduced susceptibility to tigecycline and colistin. A range of resistance profiles and associated genes were identified here, highlighting the applicability of combining genome sequencing into a routine surveillance framework to study environmental AMR as part of a wider surveillance system. Numerous potentially pathogenic strains were identified during this study. In particular, ST10, ST38, ST155, ST34, ST131, ST295, ST746 were all identified here (Supplementary File 3). These sequence types encompass both environmental and clinically associated strains. They are frequently recovered from food and human intestinal samples and have strong associations with uropathogenic and diarrhoeal disease. Recently a clinical molecular epidemiological review of ST prevalence in extra-intestinal pathogenic *E. coli* (ExPEC) isolates from Saudi Arabia found ST131 to be the dominant form observed (Alqasim, 2020). ST10, ST38 and ST155 isolates are also found commonly in the gut of humans and animals (Matamoros *et al*., 2017, Chattaway *et al*,. 2014, Castellenos *et al*., 2017). The utility of WGS to detect and further characterise strains displaying unusual or clinically relevant resistances (e.g. ESBL producing strains, isolates resistant to colistin and/or tigecycline) as demonstrated here, and in a rapid and cost-effective manner may allow genomic approaches to be further implemented as part of AMR surveillance initiatives in the future (EFSA, 2019).

To our knowledge this study represents the first attempt to design and implement a regional transnational surveillance programme for studying AMR in marine and coastal environments. It follows on from discussion/opinion articles that highlighted the need for regional coordination to tackle the emergence of AMR in the GCC region (Le Quesne *et al*., 2018, Balkhy *et al*., 2016). Both papers specifically outlined the urgent need to ascertain AMR data from environmental sources and to develop rapid baseline field surveys for the presence of AMR bacteria and associated genes in marine and aquatic environments across the GCC as part of a wider and coordinated monitoring. One of the main sources of AMR bacteria into the wider marine environment is via sewage discharge either from sewage treatment plants (STPs) or direct inputs (Aarestrup and Woolhouse 2020). In regions such as South-East Asia it is still considered that transmission via contact with contaminated environments (e.g. water and soil), and from companion or livestock animals is regarded as low risk when compared to direct human to human transmission or contact in a clinical setting (Ng & Gin 2019). However, in water scarce regions such as the GCC there is a greater reliance on water re-use for environmental enrichment (recreational parks etc), and this route as a source of exposure back to humans is often over-looked. Evidence suggests that while water treatment (including UV and chlorination) significantly reduces the concentration of bacteria in processed compared to raw effluent, the AMR of surviving bacteria in the final effluent may actually be higher than in pre or partially treated effluent (Al Jasim *et al*., 2015; Aslan *et al*., 2018). More information is needed on removal efficiencies of technology employed at STPs across the GCC when developing new AMR surveillance programmes, especially in effluents intended for discharge into the environment or reuse for irrigation (Al Jasim *et al*., 2015). Further information is also needed to better understand the potential risks to humans who use coastal waters for recreational purposes (e.g. swimming, fishing and boating). There is obviously a direct exposure risk via this route and the threat of exposure to a wide range of illness when in contact with bacterially contaminated recreation waters is clearly established, even in high-income countries with well-developed waste-water treatment infrastructure (Leonard *et al*., 2018a). Data from European coastal waters has also shown that members of the public who regularly engage in water sports are at greater risk of being exposed to, and colonised by, clinically important antibiotic-resistant *E. coli* (Leonard *et al*., 2018b). Therefore, it will be essential to further address these pathways to fully understand the risk posed by antibiotic resistant bacteria and antibiotic resistant genes on AMR prevalence in these receiving environments.

This work highlighted the need for establishing standardised methods for collection, analysis and interpretation of results. In partnership with GCC regional partners in Bahrain, Oman, Kuwait, and the United Arab Emirates, we were able to imbed and implement a coordinated environmental study that provides AMR data - albeit in a temporally and spatially constrained context - that can be utilised for various purposes, such as studying the emergence of resistant and clinically-relevant resistances, to identify new and emergent resistance genes, characterise potentially pathogenic strains present in the environment, and to suggest a comparison with AMR data generated from clinical strains to known prescription/antibiotic usage data, among others. A more structured sampling programme in future environmental surveillance designs may be better able to attribute source of *E. coli* and clinical comparison. Many AMR national action plans have yet to fully adopt a One Health approach to include environmental components of AMR. Intervention strategies can only be devised to prevent and control the spread of AMR when all possible sources and sinks have been identified. It is anticipated that significant benefits will be seen across the Gulf States if they also implement comprehensive One Health based AMR and AMU management processes, particularly if these are harmonised at a regional level. Taken together, this study demonstrates that a monitoring framework can be achieved in both a relatively short space of time and with limited infrastructure or established laboratory networks in place. In addition, this study has demonstrated greater consistency and standardization of the data collected across the region, forged transnational and international collaborations, and helped encourage improved data linkage in the GCC region.

The overall objective of the study – to design and carry out a baseline study to get an estimate of AMR in the marine environment of the Gulf – has been achieved. This has highlighted the feasibility of developing and implementing harmonised marine environmental AMR surveillance and monitoring programmes, that could directly support integrated surveillance activities across these different sectors. Key knowledge gaps remain, but the data provided here will help facilitate the development of more comprehensive action plans, and engagement with stakeholders that typically do not form the focus of existing regional and national working groups. As such it will enable a strategic overview of the coverage of different surveillance and research activities in relation to the marine and aquatic environments to help guide the development of future monitoring and research programmes.

## Data Availability

All data produced in the present work are contained in the manuscript

https://www.ncbi.nlm.nih.gov/bioproject/PRJNA691754

## Data availability statement

The assembled genomes, as well as the raw sequencing data, have been deposited at DDBJ/ENA/GenBank under the BioProject accession number PRJNA691754.

## Supplementary Files

**Supplementary file 1**. A copy of the study plan for the collection and initial processing of water samples.

**Supplementary file 2**. Data gathered during sample collection and initial sample processing e.g. location, water temperature, comments, *E. coli* concentration cfu/mL.

**Supplementary file 3**. Raw data including sample ID, location, ST type, MIC values.

**Supplementary file 4**. Statistics describing the genome assemblies of each *Escherichia coli* isolate.

**Supplementary file 5**. Frequency gene observations/relevant chromosomal mutations for resistance phenotypes.

**Supplementary file 6**. Correlation of whole genome sequencing, ECOFF values and test performances for *Escherichia coli* isolates. G+: gene/SNP present; G-: gene/SNP absent; NPV: negative predictive value; P+: phenotype resistant; P-: phenotype sensitive; PPV: positive predictive value; kappa: Cohen’s kappa coefficient.

