## Supplementary File 1 for "Establishing a marine monitoring programme to assess antibiotic resistance: a case study from the Gulf Cooperation Council (GCC) region"

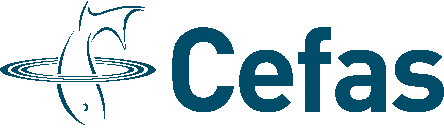

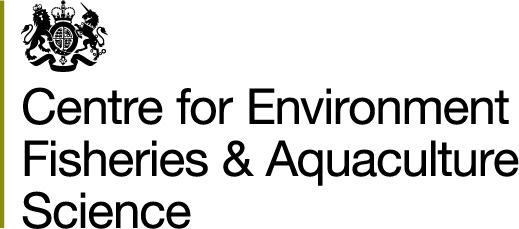

### www.cefas.co.uk

### GCC-wide rapid baseline survey of the presence of antibiotic resistant Enterobacteriaceae in the marine environment

### Study plan for sampling and processing in partner country laboratories

##### **Principle**

Organisms present in marine water are processed through filtration and entrapped on a membrane filter. The filter is placed onto tryptone bile glucuronide agar (TBX), an agar medium containing tryptone, bile salts and the chromogenic substrate 5-bromo-4-chloro-3-indolyl-β-D-glucuronide (BCIG), either as the cyclohexylammonium or sodium salt. Organisms which produce β-glucuronidase form blue colonies after incubation at 30°C for 4 hours followed by incubation at 44°C for 14 hours. These are regarded as *E. coli*. Most strains of *E. coli* express β-glucuronidase, as do some strains of Shigella and Salmonella.

##### **Limitations**

Enumeration of colonies by this method will exclude a proportion of strains of *E. coli* that are unable to grow at 44°C or that cannot express β-glucuronidase activity. Also, some proportion of *E. coli* that are stressed in marine waters cannot be recovered from environmental samples.

The method is suitable for most types of aqueous samples except those with high turbidities, which tend to block the membrane filter. This will limit the volume of sample that can be filtered. Accumulated deposit on the membrane filter may mask or inhibit the growth of bacteria. The ideal number of colonies that should be counted from a single membrane filter is approximately 20 – 80 with a maximum of 100. Samples containing more than 100 blue colonies on the filters or where it is not possible to count individual colonies should be diluted appropriately in PBS/quarter strength Ringers and the diluted sample filtered.

#### **Equipment**

- Sterile 500 ml plastic sample containers.
- Probe for measuring sea water temperature and salinity.
- Cool box with ice packs.
- Refrigerator.
- Filtration apparatus; sterile filter funnels or funnels that can be sterilised, and a vacuum filtration device.
- Cellulose nitrate filters; white, grid, 47 mm diameter, pore size 0.45 µm (supplied by Cefas).
- Smooth tipped forceps.
- Sterile quarter strength Ringers solution (tablets can be supplied by Cefas, to be prepared in country. Dissolve 1 tablet in 500 ml deionised water and sterilise by autoclaving at 121 °C for 15 minutes).
- TBX plates; store between 2 – 8 °C in the dark (supplied by Cefas).
- Nutrient agar slants; store between 2 – 25 °C (supplied by Cefas).
- Incubator; both 30 °C and 44 °C, or one cyclic incubator which can be programmed to incubate for 4 hours at 30 °C followed by 14 hours at 44 °C.

#### **Health and safety**

Local guidelines should be followed for working in the field – taking water samples.

Local risk assessments should be followed for working in a microbiology laboratory.

There are no hazards associated with TBX pre-prepared plates or nutrient agar slants. MSDS sheets will accompany the shipment.

This incubation method should not select for toxigenic *E. coli* (biohazard level 3), enabling work to be carried out in laboratories having category 2 safety facilities.

#### **Study Design**

The study design encompasses sampling, filtration, isolate selection, packaging and return to Cefas. Ideally partners will be able to fulfil all aspects of the study plan. However, if resources preclude isolate selection the original plates produced following filtration can be packaged and returned to Cefas.

#### **Sample collection**

- Print off and complete (as much as possible) a separate recording sheet for each location.
- Select 3 locations from which to collect water samples – 2 highly impacted locations near to (any) known sewage inputs, and 1 sampling location in an area used for recreational activity, but close to potential sewage inputs. Each location to be sampled once only.
- Record the water temperature, salinity, and any other observations on the recording sheet.
- Collect the water into sterile plastic bottles. Hold the bottle close to the base and submerge to a depth of about 15 cm. Collect at least 200 ml, leaving room at the top for mixing. Avoid surface scum. Tightly close the bottle and shake to homogenize the sample.
- Samples should be kept at < 10 °C between collection and processing.

##### **Sample Processing**

- Remove the TBX plates and quarter strength Ringers solution from the fridge. The TBX plates should either be left at room temperature for 2 hours, or placed at 37 °C for 30 minutes, prior to use.
- Prepare a dilution series with the water samples from 10^0^, 10^-1^, 10^-2^, 10^-3,^ 10^-4^, 10^-5^ in quarter strength Ringer’s solution.
- Set up the sterile filtration apparatus and connect to a vacuum source. Using sterile forceps place a sterile membrane filter, grid-side up, onto the porous disc of the filter base.
- Pour or pipette 10 ml of the 10^-5^ dilution into the funnel. Apply the vacuum, not exceeding 65 kPa, and filter the sample slowly through the membrane filter. Switch off the vacuum.
- Remove the funnel and transfer the membrane filter carefully onto a TBX plate ensuring that no bubbles are trapped underneath the filter.
- Using the same funnel repeat with 10 ml of the 10^-4^, 10^-3^, 10^-2^, 10^-1^ and neat (10^0^) samples, in that order.
- A sterile funnel must be used for each location if processing samples from the different locations at the same time. This can either be a separate sterile funnel, or the same funnel sterilised by placing in a boiling water bath for at least 5 minutes and allowing to cool before use.
- The time between filtration and incubation should be as short as possible, and no longer than 2 hours.
- Plates should be incubated, inverted, at 30 °C for 4 ± 0.25 hours then transferred to an incubator at 44 ± 0.5 °C and incubated for 21 ± 3 hours.

##### **Isolate selection**

- Examine the TBX membrane filters under good light. Select a dilution which contains between 20 and 100 colonies.
- Count the blue colonies (β-glucuronidase positive) see figures 1 and 2. These are regarded as *E. coli*. The combination of media selectivity, incubation temperature and the specificity of β-glucuronidase are sufficient for most practical confirmatory purposes.

Figure 1. Blue colonies of *E. coli* from a surface water on tryptone bile glucuronide agar

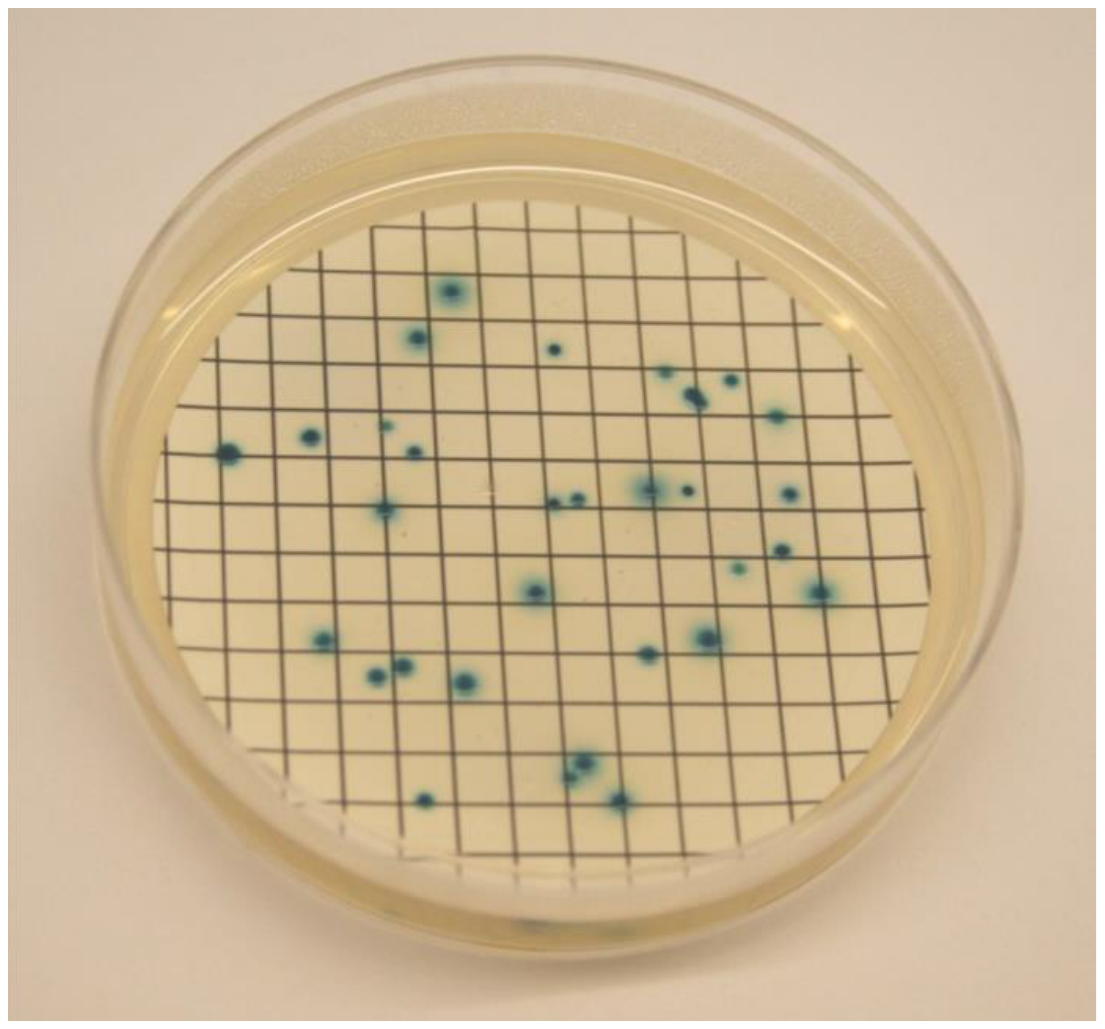

Figure 2. Blue colonies of *E. coli* from a reference culture (NCTC 09001) on tryptone bile glucuronide agar

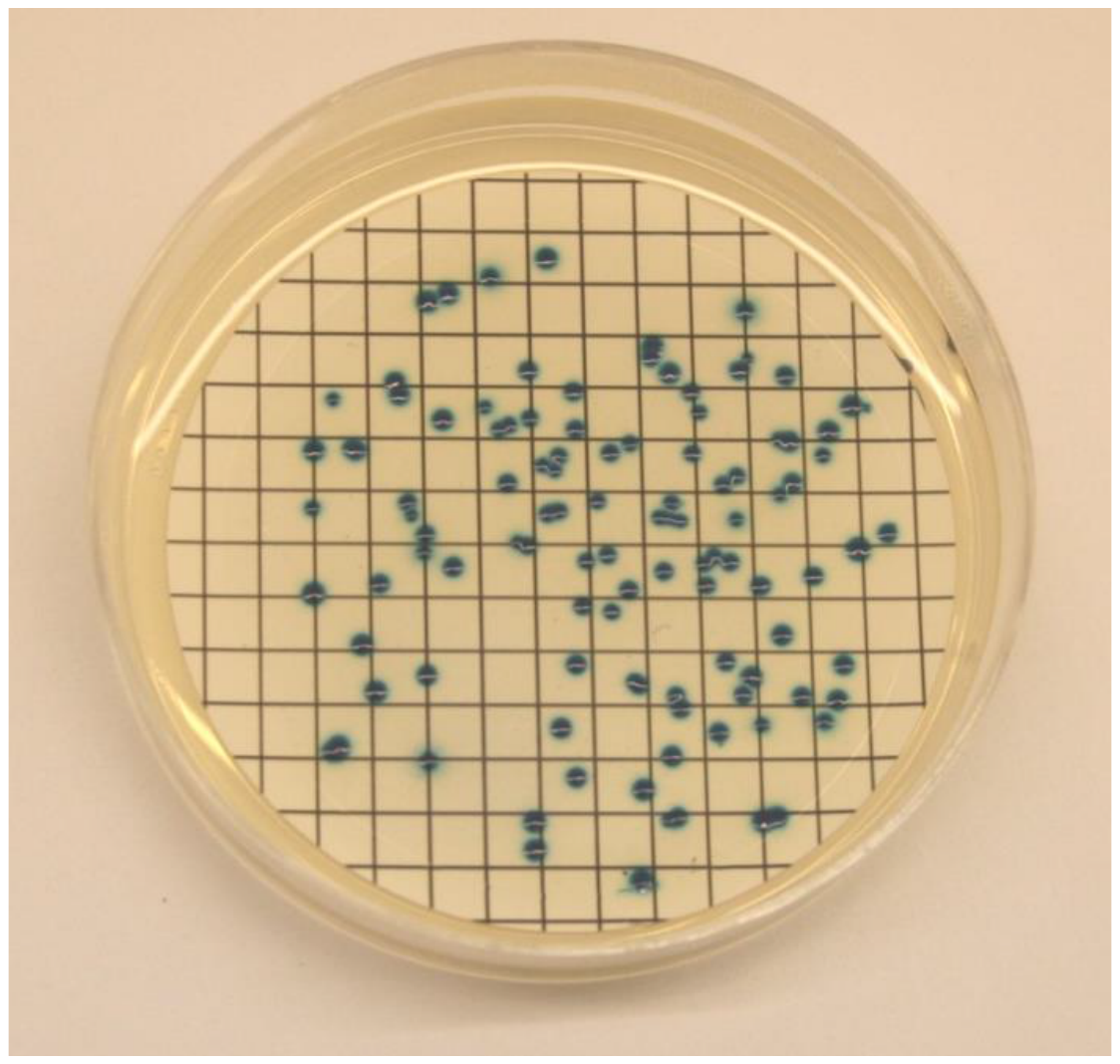

- Calculate the amount of *E. coli* in the sample. The number of *E. coli* is generally expressed as the number of colonies per 100 ml of sample. Calculate the presumptive count as follows:

Confirmed count per 100 ml =	N x 100 x DF

Volume of sample filtered (ml)

where N is the number of blue colonies counted on the TBX membrane filter, and DF is the appropriate dilution, if required.

- Pick off at least 50 blue colonies from each location. Please return up to 200 isolates from each participating country.
- Inoculate into agar slants and incubate overnight at 44 °C.
- Store the slants at 5 ± 3 °C until dispatch to Cefas.
