## Supplementary File 2 for "Establishing a marine monitoring programme to assess antibiotic resistance: a case study from the Gulf Cooperation Council (GCC) region"

| **Country** | **Location** | **Lat Long** | **Date** | **Water temp °C** | **pH** | **Salinity ppt** | **Comment** | **Incubation conditions** | **Total E. coli**  **cfu/ml** | **No. of E. coli isolates tested** |
| --- | --- | --- | --- | --- | --- | --- | --- | --- | --- | --- |
| Kuwait | AlGazali | 29°347784 N  47°911974 E | 17/12/2018 | 14.0 | 7.9 | 40.0 | Obvious sewage. Effluent. Bad smell | 30 °C 3 hrs  44 °C 20 hrs | 5.0 x 10^5^ | 100 |
|  | Alsalam | 29°357207 N  47°946784 E | 17/12/2018 | 14.0 | 8.1 | 41.0 | Obvious sewage. Effluent. Bad smell | 30 °C 3 hrs  44 °C 20 hrs | 2.8 x 10^5^ | 100 |
|  | Doha Bay | 29°369112 N  47°742780 E | 15/01/2019 | 14.0 | 7.6 | 41.0 | Obvious sewage. Effluent. Bad smell | 30 °C 3 hrs  44 °C 20 hrs | 3.5 x 10^5^ | 23 |
| Bahrain | Askar | 26°03.046 N  50°37.288 E | 18/12/2018 | 21.5 | 7.78 | 42.57 | 30 metres from a small sewage outlet. Effluent flowing while sample was being taken. | 30 °C 4 hrs  44 °C 21.5 hrs | 6.2 x 10^2^ | 52 |
|  | Nabih Saleh | 26°10.830 N  50°34.807 E | 18/12/2018 | 18.7 | 7.92 | 35.4 | Coastal area where a few fishing boats are kept. No swimming activities. | 30 °C 4 hrs  44 °C 21.5 hrs | 1.9 x 10^3^ | 52 |
|  | Tubli Bay | 26°11.808 N  50°34.112 E | 18/12/2018 | 23.0 | 7.41 | 34.92 | 400 metres from a large-scale sewage treatment plant discharge. Low tide, water less than 1 metre deep. Directly affected by sewage discharge. | 30 °C 4 hrs  44 °C 21.5 hrs | 2.3 x 10^5^ | 52 |
| Oman | Al Qurum | 23°38’01.6’ N  58°29’11.2 E | 12/02/2019 | 21.0 | 7.03 | Not tested | Approx. 200 m from the beach and a sewage pipe which collects waste from nearby houses | 44°C 21 hours | 1.8 x 10^6^ | 21 |
|  | Darsait | 23°37’03.6 N  58°32’26.5 E | 12/02/2019 | 22.0 | 8.28 | Not tested | Sewage water collected under the road mixed with rainwater going to the wadi under the road. Looked like clear tap water. | 44°C 21 hours | 3.2 x 10^2^ | 47 |
|  | Muttrah | 23°37’30.9 N  58°33’47.4 E | 12/02/2019 | 27.0 | 7.11 | Not tested | Water collected from the surface of sewage (sink) near to the suq. Very dirty. | 44°C 21 hours | 9.6 x 10^3^ | 34 |
|  | Al Qurum | 23°38’01.6’ N  58°29’11.2 E | 29/04/2019 | 27.0 | 7.13 | Not tested | Approx. 200 m from the beach and a sewage pipe which collects waste from nearby houses. | 44°C 21 hours | 7.0 x 10^6^ | 50 |
| UAE | Dubai Creek Dhow Wharfage 1 | 25°258345 N  55°316281 E | 15/05/2019 | 29.4 | 8.38 | 39.7 | Dhow Wharfage where Dhow boats berth and load/unload shipment, quite dirty water. | 30°C 4 hours  44°C 17 hours | 1.7 x 10^2^ | 20 |
|  | Dubai Creek Dhow Wharfage 2 | 25°257273 N  55°317837 E | 11/02/2019 | 22.8 | 8.13 | 39.9 | Dhow Wharfage where Dhow boats berth and load/unload shipment, quite dirty water. | 30°C 4 hours  44°C 17 hours | *unknown | 6 |
|  | Dubai Creek  Ras Al Khor | 25°211748 N  55°347388 E | 11/02/2019 | 22.9 | 8.35 | 37.0 | Upper part of Dubai Creek, fairly close to outlet of tertiary treated water from Al Awir STP, impacted by Ras Al Khor mangrove swamp, close to building site, murky brown water | 30°C 4 hours  44°C 17 hours | *unknown | 2 |
|  | Dubai Creek Wharfage 3 | Not available | 15/05/2019 | 29.4 | 8.4 | 39.9 | Dhow Wharfage where Dhow boats berth and load/unload shipment, quite dirty water. | 30°C 4 hours  44°C 17 hours | 1 x 10^1^ | 1 |

* Few individual colonies removed from the filter. Smear prevented accurate enumeration.
